# Mapping the distribution and describing the first cases from an ongoing outbreak of a New Strain of mpox in South Kivu, Eastern Democratic Republic of Congo between September 2023 to April 2024

**DOI:** 10.1101/2024.05.10.24307057

**Authors:** Leandre Murhula Masirika, David F. Nieuwenhuijse, Pacifique Ndishimye, Jean Claude Udahemuka, Bilembo Kitwanda Steeven, Nzigire Barhatwira Gisèle, Jean Pierre Musabyimana, Baganda Ntahuma Daniel, Théophile Kiluba wa Kiluba, Franklin Kumbana Mweshi, Polepole Ngabo, Theophile Tambala, Mazambi Mambo Divin, Bahati Mutalemba Chance, Léandre Mutimbwa Mambo, Leonard Schuele, Justin Bengehya Mbiribindi, Gustavo Sganzerla Martinez, David J Kelvin, Gaston Lubambo Maboko, Bas B. Oude Munnink, Trudie Lang, Frank M. Aarestrup, Christian Gortazar, Marion Koopmans, Freddy Belesi Siangoli

## Abstract

**Background:** In September 2023, an outbreak of mpox was reported in the eastern part, South Kivu Province, of Democratic Republic of the Congo. This paper aims to provide a summary from several ongoing and completed studies to share initial insights into the time trend and spatial distribution, the links we have observed between level of mpox cases with population density and the presence of professional sex workers (PSW) in bars within affected heath areas of Kamituga health zone. We also seek to share our initial observations on the novel characteristics of this new and emergent strain.

**Methods:** Consenting patients admitted with mpox-like symptoms to the Kamituga hospital were recruited to an observational cohort study to understand the clinical characteristics and household perceptions and treatment seeking behaviours. We mapped the demographic and clinical characteristics of all individuals between September 2023 and April 2024.

**Findings:** 371 (suspected) mpox cases were admitted to the Kamituga hospital. There were slightly more female than male cases (192/371 [52%] versus 179/371, [48%], and cases were reported from 15 health areas. The majority of cases were reported in Mero (115/371 [31%]), followed by Kimbangu (65/371[18%]), Kabukungu (63/371[17%]), Asuku (47/371 [13%]), Soluluyu (19/371 [5,12%]), Katunga (17/371 [4,58%]), Kalingi, Poudriere (12/371 [3,23%]) respectively, and Poly Afia (7/371 [1,89%]). During this period, 4 deaths occurred and 4 out of 8 women who were pregnant had fetal loss . Three healthcare workers acquired infection when caring for patients. In depth case ascertainment showed that 88,4 % of patients reported recent visits to bars for (professional) sexual interactions as a likely source of infection. Our findings suggest that the epidemic in South Kivu is driven by transmission in a network of professional sex workers, working in bars. The continued spread of the mpox virus in Kamituga health zone and other health zones of South -Kivu points at a critical need for cross border surveillance, and the potential for rapid vaccination of sex workers as potential intervention.

**Interpretation:** These data suggest that the rapid transmission of mpox virus is related to interactions with professional sex workers (PSW) in bars within densely populated health areas. The expanding number of cases and the recent spillover to 16 other nearby health zones of South -Kivu including a health district bordering Rwanda and Burundi stresses the need for cross border surveillance. In addition, enhanced response action is needed, including vaccination health education programmes to limit further escalation and stop this outbreak.

## INTRODUCTION

Human mpox disease (formerly known as monkeypox) is an emerging zoonotic disease caused by the monkeypox virus (MPXV). MPXV is a double-stranded DNA virus that belongs to the genus *Orthopoxvirus* within the *Poxviridae* family, subfamily *Chordopoxvirinae*. The first recorded human case of MPXV was in 1970 when a nine-month-old child was admitted to Basankusu Hospital in the Democratic Republic of Congo (DRC). The child suffered from a disease similar to smallpox, from which a virus similar to MPXV was isolated [1,2]. The clinical manifestations associated with MPXV resemble those of smallpox, a disease which has been globally eradicated since 1980. Infection in humans manifests as fever, swollen lymph nodes, and fatigue followed by a rash (genital, anorectal, or oral), with macular lesions progressing to papules, vesicles, pustules, and scabs, usually on the face, hands, and feet for two to five weeks [4,6,16].

MPXV is historically enzootic in Western and Central Africa and there have been sporadic mpox outbreaks in humans with increasing frequency primarily in West and Central African countries. Though the exact host reservoir for monkeypox is still unknown, some studies have shown that the reservoir is likely to be one or more species of rodents or squirrels that inhabit the secondary forest of central Africa [3,5,20]. Mpox achieved global attention after 2003 due to the first outbreak in the USA linked to infected pet prairie dogs [3]. Wild-caught native prairie dogs co-housed with rodents imported from Ghana in Western Africa in a large animal trading facility and subsequently sold as pets were thought to be the primary source of the outbreak [1,3]. A total of 81 human cases were reported, of which 31 were laboratory confirmed. A subsequent serological study suggested there may have been underdiagnosis [16].

Since 2003, several cases of mpox have been reported in various countries. A study conducted between November 2005 and 2007 in DRC concluded that the incidence of human infections had massively increased since the cessation of smallpox vaccination programs, in line with a growing immunologically naïeve proportion of the population. The study also mapped increased risk to forested areas with an enzootic reservoir [11].

The DRC is the sole country to have been continually reporting mpox cases in the last five decades, but there was limited global interest in supporting containment efforts.

This changed when a widespread outbreak of mpox occurred worldwide in May 2022, when the first case was confirmed in the United Kingdom in a man traveling from Nigeria [14]. This ongoing outbreak, caused by a lineage II MPXV, is primarily transmitted through networks of men engaging in frequent unprotected sexual activities with other men (MSM) [14,15]. Although previous mpox outbreaks in the regions with enzootic MPVX reservoirs have suggested potential for spread to more diverse populations and through different modes of transmission, this has been very limited in the global outbreak.

Two genetically distinct clades of MPVX have been described: Clade I (formerly known as the Congo Basin or Central African clade) and Clade II (formerly known as the West African clade). Since August 2022, Clade II has been subdivided into two subclades, Clade IIa and Clade IIb. Viruses from Clade I and Clade II have a nucleotide sequence similarity of 99.4% [10]. MPXV Clade I is thought to cause more severe disease with a case fatality rate (CFR) of up to 10.6%, while MPXV Clade II is associated with milder symptoms and a lower CFR of roughly 0.1% [10,17,19]. Clade I is typically associated with some (limited) transmission within households and often identifiable to a source of infection from bushmeat, with previously no associations with sexual transmission. Since 2022, Clade II infections are primarily sexually transmitted.

In the DRC, numbers of people with suspected infection with MPXV have increased since the start of 2023. A total of 12,569 suspected mpox cases have been reported up to November 12th 2023, the highest number of annual cases ever recorded in the country. The case fatality rate has been estimated to be 4.6% [11]. New cases have occurred in remote geographical areas of the country where the disease was previously not observed, including South Kivu province which is far from other cases. Recently, we showed that the outbreak in South Kivu is associated with a novel sub-lineage of clade I viruses [6,7]. The majority of these cases seem to be transmitted through (hetero)sexual contact, with some person-to-person transmission within and outside the households.

Here, we provide a detailed overview of the evolution of the outbreak and spatial distribution of cases in South Kivu from September 29^th^ 2023 to April 21^st,^ 2024, in relation to population density and density of bars with sex workers. We also report the emerging disease characteristic of this new strain because we want to convey these findings rapidly for public health importance.

## METHODS

### Study design and participants

Patients are being admitted routinely to Kamituga Hospital during this ongoing mpox outbreak. All patients are invited to take part in the ongoing research studies that are seeking to address the many unknowns that are becoming clear in the different presentation and transmission of this new strain. Throught these mixed-methods prospective observational cohort studies, we have collected the demographic and clinical characteristic data of all individuals with MPVX infection symptoms admitted at the Kamituga Hospital between September 2023 and April 2024 who agreed to participate in the study. We also have also geolocated these cases from the notification database in the health zones of South-Kivu province. The ethical clearance to conduct these studies was obtained from the Ethical Review Committee of the Catholic University of Bukavu (Number UCB/CIES/NC/022/2023 and University of Oxford : OxTREC Reference : 517-24). All study participants were introduced to the observational study and given the option to participate by providing informed consent or, in the case that the participant was a minor, through parental permission. Patient information was anonymized and we confirm the IDs we used make the study participants unidentifiable.

### Procedures and sample collection

Routine data regarding age, gender, professional occupation, clinical symptoms, geolocations of mpox case and concomitant presence of sexually transmitted infections (STIs), and comorbidities were collected from patient records (or hospital investigation forms) and entered into a secured, anonymized database. The study team was informed about the objectives of the study, the consent process, and the importance of standardised reporting

Admission to the Kamituga Hospital was based upon clinical diagnosis of human MPVX infection by hospital staff. A <u>confirmed</u> MPVX case was defined as an individual with laboratory-confirmed infection. A case was listed as “<u>suspected</u>” if a patient had an acute illness with fever, intense headache, myalgia, and back pain, followed by one to three days of a progressively developing rash often starting on the face and spreading on the body. Finally, a case was listed as “<u>probable</u>” if it satisfied the clinical definitions of suspected cases and had an epidemiological link to a confirmed or probable case but was not laboratory-confirmed. The suspected cases were PCR tested in the National Institute for Biomedical fot Research I (INRB). A skin lesion was defined as a single circumscribed area and included presentations comparable to papules, pustules, fluid-filled vesicles, or eschars.

### Mapping of mpox cases

Geographic and epidemiological data was processed using the R programming language. The epidemiological curve was plotted using the “ggplot2” [26] package. For the geographical maps cases and sexworkers were scaled by population density and aggregated by health area defined by the shapefiles provided by national health ministry in Democratic Republic of the Congo available online: https://data.humdata.org/dataset/drc-health-data. Map plots were made by using the “sf” [27] and “ggmap” [28] R packages.

## RESULTS

### Study area

Kamituga is South Kivu’s largest gold mining city located in the territory of Mwenga, part of the South Kivu province. The South Kivu province borders in the north with the North Kivu province, south and west with the Maniema province, and south with the Tanganyika province. To the east, South Kivu borders Rwanda, Burundi, and Tanzania. The city of Kamituga has more than 241,642 inhabitants (based on the 2024 Kamituga Health Zone report), of which ∼20,000 are employed in the mining industry. The remaining residents depend directly or indirectly on artisanal and small-scale gold mining (ASGM) for their livelihood.

### Epidemiological curve plot and maps of the outbreak evolution in time and space within health areas of Kamituga

A total of three hundred and seventy-one hospitalized mpox case records were listed as confirmed, probable or suspected (**Figure 1 A**) between September 29^th^, 2023 and April 21^st^, 2024. Three of these were health care workers, four deaths occured (1%) and four abortions were reported. In contrast to other outbreaks in the Democratic Republic of Congo, only 25 % (79/371) of suspected cases were children under 15 years of age and the majority of patients were aged between 16 -26 years old.

The highest number of admissions in a single week for the year 2023, i.e., 14, was reported in weeks 43, 48, and 52. In 2024, the number of mpox cases sharply increased with the highest number of 32 admissions reported in week 16. The 4 week moving average also increased from around 10 hospitalized cases per week in 2023 with a gradual increase to over 20 cases hospitalized per week in the Kamituga hospital since the beginning of 2024 as reported in (**Figure 1 A, Figure 1B**). The outbreak expanded to Bigombe, Luliba, Isopo, Kele Katutu, and Kankanga respectively in the epidemiological weeks 7, 10, 11, and 15 of the year 2024.

**Figure 1.**
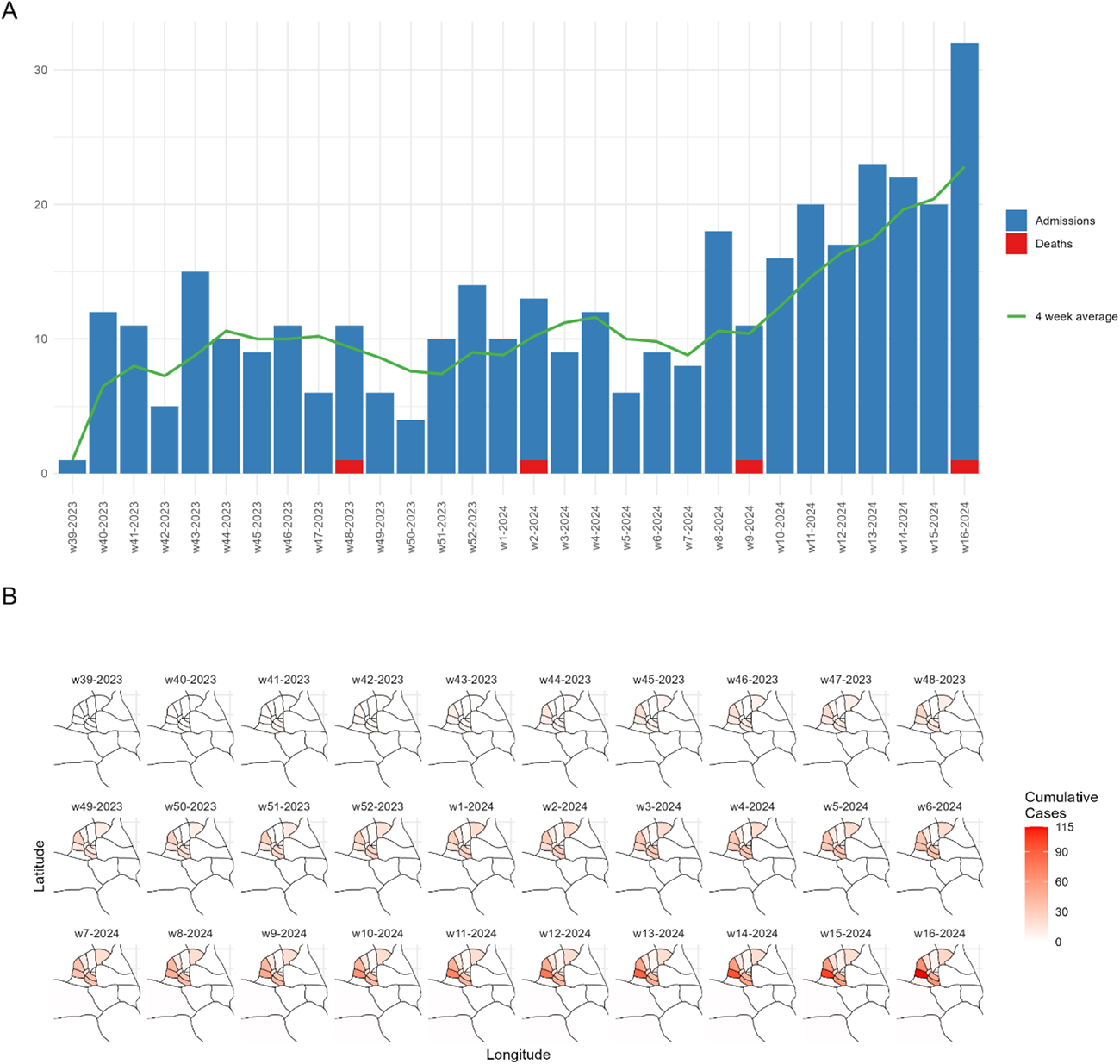
**A**: Epicurve of outbreak based on Kamituga health zone surveillance dataset for the mpox patients’ admissions at Kamituga hospital and the deaths per epidemiological week between September 29^th^, 2023 to April 21^st^, 2024. **B**: Map of the epi curve of the outbreak showing evolution in time and space within health areas of Kamituga health zone

### Description of fatal cases

Kamituga health zone is the only health zone in South Kivu continually reporting mpox cases and considered the mpox epicenter since September 2023. During this period, 4 deaths have occurred. The first case of death was reported in November of 2023, a woman whose age ranged between 26-30 years came in (**A1)** health area. She was first admitted to a health center located in (**A1)** health area but transferred to Kamituga hospital with an antecedent of cesarean section she underwent two weeks earlier. She presented mpox like symptoms like fever, rash, dysphagia, adenopathy, genital lesions followed by belly bloating and intensive muscle pain. An acute peritonitis was diagnosed. In addition, she was tested positive by mpox PCR test while an HIV test was negative. Treatment with ceftriaxone, cloxacillin, tramadol and metronidazole intravenous were administered for 7 to 10 days afterwards. She underwent a surgical operation from which she did not recover.

A second death was reported in Junuary 2024, the patients’s age ranged betwwen 21-25 from **(A2)** health area. She arrived at the hospital with mpox-like symptoms. MPVX and HIV were diagnosed, and the patient was treated with ceftriaxone, cloxacillin, tramadol and metronidazole intravenous and ibuprofen, vitamin C were administered for 7 to10 days. The patient developed convulsions and then died.

A third death was reported in March, 2024. Again, the patient of interval age was between 21-25-year-old from (**A3)** health area who arrived at the hospital with mpox like symptoms. HIV was tested negative. Treatment like ceftriaxone, cloxacillin, tramadol and metronidazole intravenously were administered for 7 to 10 days. Complications occurred when instead of the pustules falling into crusts, for this patient the pustules burst into opened wounds with stench. The medical staff attributed this to a possible bacterial superinfection of bacteria C*lostridium tetani* and she died.

The fourth death was a patient who came from **(A4**) heath area and whose age ranged between 26-30 years old. The patient arrived at the hospital in April, 2024 with severe symptoms. He was tested HIV negative, and was treated with cloxacillin, ceftriaxone, tramadol, and paracetamol for 7 to 10 days. He died from respiratory distress while PCR was pending.

### Further observations of compications

During the ongoing mpox outbreak in Kamituga health zone, 8 pregnant women were admitted to the hospital, 4 of which aborted. The first and second cases were women both ranged between 26-30 years old from the health areas **(A2**)and (**A5)**, respectively. One was admitted in November 2023 and another in January 2024 for severe mpox symptoms at the first quarter of pregnancy. Both of them tested positive by PCR and negative for HIV. Examination of echography revealed an absence of fetal movement and both women aborted.

The third abortion case was a wamon ranged between 26-30 interval age from (**A2)** health area, admitted to the hospital on April, 2024 for severe mpox symptoms, at the 2nd quarter of the pregnancy. An echography examination concluded fetal death in utero. She underwent surgical intervention for cervical dystocia. The medical staff observed a third degree macerated fetus with skin rashes similar to those of mpox. HIV was tested positive with the MPVX PCR still pending.

The fourth abortion was a woman whose age ranged between 20-25 years old from (**A6)** health area who was admitted to the hospital on March, 2024 with severe mpox symptoms. After 21 days of hospitalization she aborted at the 1st quarter of the pregnancy. Treatment used were ceftriaxone, promethazine, tramadol and metronidazole intravenously administered for 7 to 10 days. HIV was tested negative and an MPVX PCR is still pending. All of the reported pregnant women lived in the health area highly affected by mpox and all of them had contact with bar exposure.

### Sexual exposure and link to bars

Kamituga health zone has 23 official health areas. On September 29^th^ 2023, the index mpox case was reported in the Poudriere health area, and was linked to a bar visit. We interviewed hospitalized cases with a standardised case reporting form including assessing potential sexual exposures. Of the 371 hospitalised mpox cases, 88,4% (328/371) reported recent sexual contact in bars among which 47,16% (192/371) were female and 41,23% (153/371) were male. Only a few cases reported to not have sexual contact in bars: in total 11,59% (43/371) with 7,00% (26/371) males and 4,58 % (17/371) females.

### Census of Professional sex workers within bars per health area

We carried out the census of professional sex workers (PSW) in bars of Kamituga health zone. Kamituga mining city has 67 bars distributed within health areas ; Mero has 13 bars, Kabukungu 10 bars, Soluluyu 8, Poudriere 6, Poly Afia and Kimbangu 5 bars, Kalingi 4 bars, Asuku and Kibe 3 bars, and finally Bigombe, Mulambula, Kele Katutu, katunga and Luliba 2 bars.

We also inventoried the number of PSWs working in bars (751 in total) within Kamituga health zones. The highest numbers were reported in Mero health area with 21,44% (161/751) followed by Kabungu with 20,24% (152/751), Ploy Africa 10,39 % (78/751), Soluluyu 9,99% (75/751), Kalingi 7,59% (57/751), Kimbangu 5,99% (45/751), Asuku 5,06% (38/751), Kibe 4,53% (34/751), Poudriere 4,26% (32/751), Kele Katutu 2,80% (21/751), Bigombe, Luliba 2,40 % (18/751), Mulambula 2,00% (15/751), and Katunga 0,93% (7/751) **(Figure2 C-A)**

### Mpox cases reported within health areas

We mapped the health areas reporting mpox cases and bars displayed within them (Figure 2). The Health area Mero reported the highest number of mpox cases (115/371 [31%]), followed by Kimbangu (65/371[18%]), Kabukungu (63/371[17%]), Asuku (47/371 [13%]), Soluluyu (19/371 [5,12%]), Katunga (17/371 [4,58%]), Kalingi, Poudriere (12/371 [3,23%]), and Poly Afia (7/371 [1,89%]). At the beginning of 2024, the outbreak expanded to other health areas that had not been reporting mpox cases in 2023, namely Bigombe 1/371 [0,27%]) Isopo (1/371 [0,27%]), Kele Katutu (2/371[0,45%]) Kibe (1/371 [0,27%]) Lubiba (7/371 [1,89 %]), and Kankanga (2/371, [0,54%]) (**Figure 2D-B**).

### Mpox and population density of Kamituga Heath zone

We mapped the population density of the Kamituga health zone within health areas. In the 2023 Mero had the highest population 25,598/214,280 (11,95%), Kimbangu 16,132/214,280 (7,53%), Poudriere 12,931/214,280 (6,03%), Katunga 13,492 /214,280 (6,30%), Kalingi 12,888/214,280 (6,01%), Kabukungu 13,758/214,280 (6,42%) (**Figure 2E**). Our findings suggest that mpox cases from the ongoing outbreak are distributed in the health areas with higher populations and housing bars recruiting sex workers. This suggests that the outbreak of mpox in Kamituga is sustained and maintained by the sex industry through females who act as PSWs and males as barmen.

**Figure 2:**
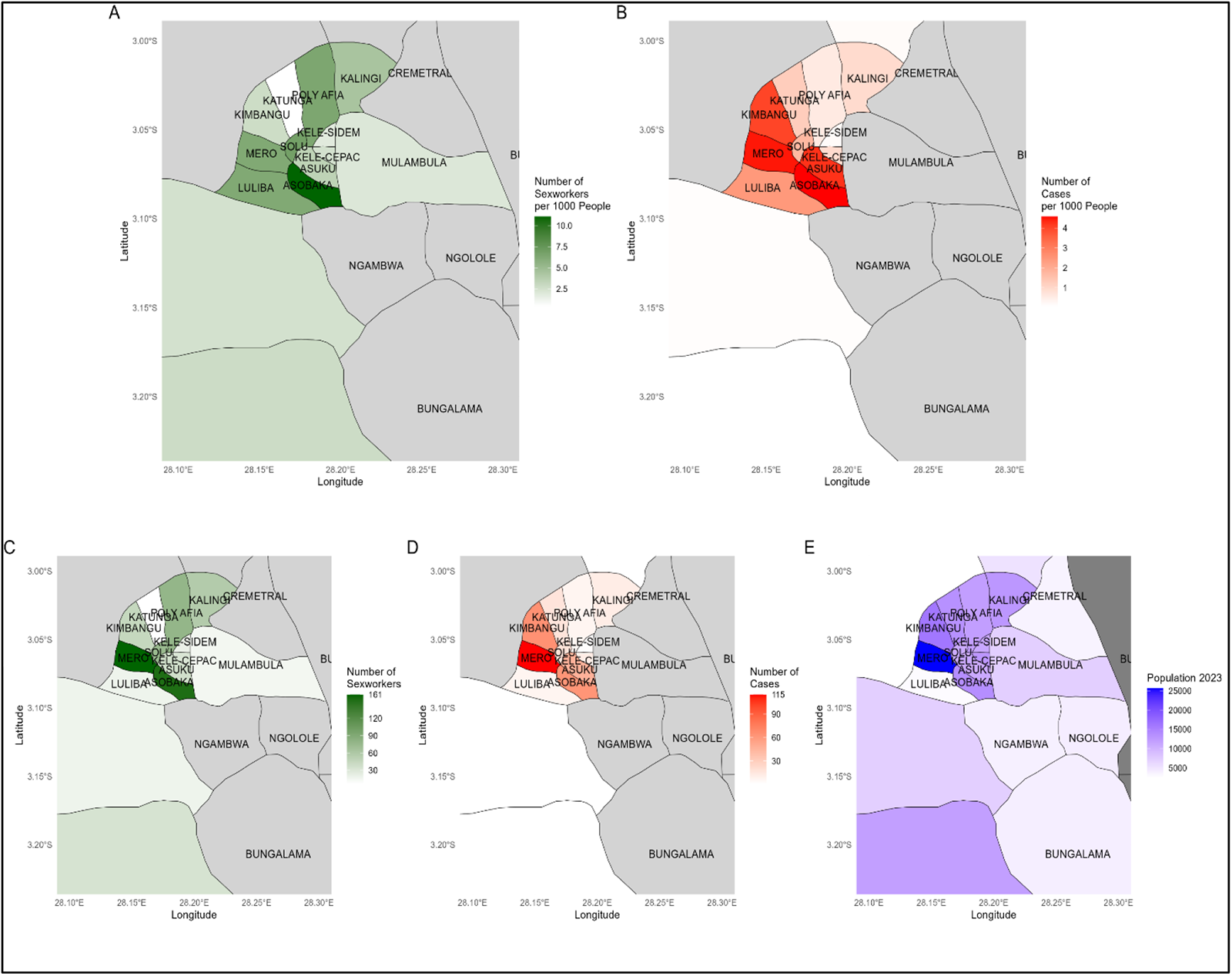
**Map(A)** reports the sex workers per 1000 inhabitants, **Map(B)** reports the numbers mpox cases per 1000 inhabitants, **Map(C)** reports the number of sex workers, **Map(D**) reports the number of mpox cases, **Map(E)** reports population density (2023) per health area in Kamituga health zone, between September 29^th^, 2023 and April 21^th^, 2024, South -Kivu, DRC.

### Expansion of mpox cases to other health areas of South-Kivu during the ongoing outbreak

Besides the large and sustained outbreak of Kamituga, mpox cases were reported in other health zones in South Kivu in the same period. The additional 63 cases were distributed as follows: the health zone of Nyangezi reported the highest number of the mpox cases (28 mpox cases), Kadutu (10 mpox cases), Shabunda (10 mpox cases), Ibanda (2 mpox cases), Walungu (2 mpox cases), Mwenga (2 mpox cases), Kaniola (1 mpox cases), Fizi (1 mpox cases), Uvira (1 mpox cases), Itombwe (1 mpox cases), Kalole (1 mpox cases), Kitutu (1 mpox cases), Nyantende (1 mpox cases), Bagire (1 mpox cases) and Idjwi (1 mpox cases).

## Discussion

Few formal case investigations of Clade I have been documented during outbreaks in Central Africa, and no sexual transmission of Clade I cases was reported until April 2023. The first documented case of the sexually transmitted mpox clade I virus involved a male resident of Belgium who had traveled in the Democratic Republic of the Congo and had sexual encounters in Kwango province (southwest) [9]. Prior to 2023, South Kivu Province had not previously reported any continuous mpox cases, except for the limited sporadic mpox cases reported in Shabunda territory, South-Kivu in 2014 [8]. We recently identified a new mpox sublineage from the current outbreak in South Kivu and suggested a different route of transmission of the virus [7,25]. We showed that in this outbreak the virus spreads primarily through heterosexual contact [6].

This data we have compiled here provides additional insights into the time trend and spatial distribution of the ongoing mpox outbreak in South Kivu, Democratic Republic of the Congo (DRC), and sheds light on the links of increasing mpox cases with population density and the presence of PSWs in bars within affected heath areas.

During this outbreak, 8 pregnant women were admitted at Kamituga hospital among whom 4 aborted, supporting the potential for intrauterine transmission of MPVX in pregnant people as also previously suggested [12,13,]. In 2008 a MPVX Clade I case of fetal death after placental infection was reported in at the Kole Hospital in Kole, located in the Sankuru District of Kasaï-Oriental Province, which findings suggested risks of intrauterine infection emphasizing that clinicians should be aware of this potential complication [13].

The health areas reporting continuous mpox cases were in the cities with a high number of bars, that are sustaining the local sex industry. Since the main activity and economic driver is gold mining, the sex work industry has served as a gainful occupation for females as PSWs and for males who act as their managers. Thus, during hospital records and investigations the majority of the patients reported to have had contact with bars in the two or three weeks before they started to develop mpox-like symptoms. The highest number of mpox cases recorded at the hospital originated from the Mero health area that had a link to such bars.

The findings revealed that the majority of mpox patients reported in Kamituga lived in areas overpopulated health areas such as Mero, Kimbangu, Kabukungu, Asuku, poudriere, Poly Afia Katunga, Soluluyu and Kalingi within higher population densities and the majority of reported cases linked to bar exposure to (88,4%), maintaining sex over activities with poor hygienic conditions.

We also report that 16 health zones of South-Kivu have had mpox cases. In addition to Kamituga, the second highest number of mpox cases is reported in the mining areas of Nyangezi health zone. Nyangezi has similar activity of mining including the city of Kamanyola which is rich in cassiterite mining, and is located at the crossroads of three countries, Congo, Rwanda and Burundi. People coming from remote mining areas spend the night in Kamanyola city and possibly in bars before crossing either to Rwanda, Burundi or to other nearby towns in DRC such as Bukavu and Goma. PSWs from Burundi, DRC and Rwanda work in bars in Kamanyola bar activity, pointing at an increased risk for cross-border transmission.

Important unknowns remain and we are continuing our programme of studies. These seek to assess the relationship between transmission and severity of disease, the long-term complications and effect of this infection and also a strong component of qualitative research with these patients when they have returned home to understand their perception, behaviours and experiences. We seek this evidence within our global and collaborative team and aim to determine evidence as rapidly as possible to help understand and stop further transmission.

## Conclusion

In conclusion, we report an ongoing mpox outbreak expanded to over 15 Kamituga zone’s health areas, thus far, causing 4 deaths and 4 abortions in 317 persons admitted to the Kamituga hospital. The outbreak appears to be driven by sexual activity with PSWs linked to bars supporting the previously undocumented model of heterosexual transmission. Because of the expansion of the outbreak to bars employing PSW from neighbouring countries, cross border surveillance and access to rapid diagnostics are crucial to control further dispersal. We also suggest that risk-targeted vaccination should be a priority.

## Data Availability

clinical data is available upon the reasonable request to the correspoding auther

## Author Approval

All authors approved the final version of the manuscript.

LMM, JCU, PN, KM, TL, conceptualized and designed the study LMM, TL, KM, NBG, FKM, LS, DF,CG, PN, DFN, FMA, BBOM, contributed to data acquisition and interpretation, drafted, and cross-reviewed the manuscript. LMM, MMD,BND, BMC, FBS, JMB, TL, GLM, TKWK, KJV, GSM, TT involved in sample collection and investigation. All authors approved the final version.

## Ethical approval

The ethical clearance to conduct these studies was obtained from the Ethical Review Committee of the Catholic University of Bukavu (Number UCB/CIES/NC/022/2023 and University of Oxford : OxTREC Reference : 517-24).

## Acknowledgements

We would like to thank the Provincial Division of Health (DPS) of South-Kivu and Kamituga Health Zone (KHZ) for their collaboration during the study. We greatly thank the National Biomedical Research Institute (INRB) for mpox cases confirmation during the ongoing outbreak. We greatly thank the EU Horizon 2020 grants VEO (874735) and the Global Health EDCTP3 Joint Undertaking (Global Health EDCTP3) program under grant agreement No. 101103059 (GREATLIFE) to build local capacity in terms of sequencing. We greatly thank Global Health Network (GHN) for access mpox disease characterisation protocol training and ISARIC for sharing the mpox disease characterisation protocol.

We would like to thank the Dalhousie University, Canada Research Chair in Translational Vaccinology and Inflammation to have been the first to support the Kamituga hospital at the beginning of the outbreak. Finally, we greatly thank Wildlife Conservation Network (WCN) and Conservation Action Research Network (CARN) for the scholarship and research support they awarded to the first author.

## References

1. Von Magnus P, Andersen EK, Petersen KB, Birch-Andersen A. A pox-like disease in cynomolgus monkeys. Acta Path. Micro. Scand. 1959;46(2):156–176. [Google Scholar] Discovery of monkeypox virus.

2. Ladnyj I, Ziegler P, Kima EJBWHO. A human infection caused by monkeypox virus in Basankusu Territory, Democratic Republic of the Congo. Bull World Health Organ. 1972 ;46(5):593.

3. Curaudeau, M., Besombes, C., Nakouné, E., Fontanet, A., Gessain, A., & Hassanin, A. (2023). Identifying the Most Probable Mammal Reservoir Hosts for Monkeypox Virus Based on Ecological Niche Comparisons. Viruses, 15(3). 10.3390/v15030727

4. Kaler, J., Hussain, A., Flores, G., Kheiri, S., & Desrosiers, D. (2022). Monkeypox: A Comprehensive Review of Transmission, Pathogenesis, and Manifestation. Cureus. 10.7759/cureus.26531

5. Doty, J. B., Malekani, J. M., Kalemba, L. N., Stanley, W. T., Monroe, B. P., Nakazawa, Y. U., Mauldin, M. R., Bakambana, T. L., Dja Liyandja, T. L., Braden, Z. H., Wallace, R. M., Malekani, D. V., McCollum, A. M., Gallardo-Romero, N., Kondas, A., Townsend Peterson, A., Osorio, J. E., Rocke, T. E., Karem, K. L., … Carroll, D. S. (2017). Assessing monkeypox virus prevalence in small mammals at the human–animal interface in the democratic republic of the congo. Viruses, 9(10). 10.3390/v9100283

6. Murhula Masirika, L., Claude Udahemuka, J., Ndishimye, P., Sganzerla Martinez, G., Kelvin, P., Bubala Nadine, M., Kitwanda Steeven, B., Kumbana Mweshi, F., Mutimbwa Mambo, L., Oude Munnink, B. B., Bengehya Mbiribindi, J., Belesi Siangoli, F., Lang, T., Malekani, J. M., Aarestrup, F. M., Koopmans, M., Schuele, L., Pierre Musabvimana, J., Umutoni, B., … Flores, L. (n.d.). Epidemiology, clinical characteristics, and transmission patterns of a novel Mpox (Monkeypox) outbreak in eastern Democratic Republic of the Congo (DRC): an observational, cross-sectional cohort study. 10.1101/2024.03.05.24303395

7. Murhula Masirika, L., Claude Udahemuka, J., Schuele, L., Ndishimye, P., Otani, S., Bengehya Mbiribindi, J., Marekani, J. M., Mutimbwa Mambo, L., Malyamungu Bubala, N., Boter, M., Nieuwenhuijse, D. F., Lang, T., Balyahamwabo Kalalizi, E., Pierre Musabyimana, J., Aarestrup, F. M., Koopmans, M., Oude Munnink, B. B., Belesi Siangoli, F., Leandre Murhula, M., … Freddy Belesi, S. (n.d.). Rapid communication Ongoing mpox outbreak in Kamituga, South Kivu province, associated with monkeypox virus of a novel Clade I sub-lineage, Democratic Republic of the Congo, 2024 *Correspondence: Leandre Murhula*. 1. 10.2807/1560

8. McCollum, A. M., Nakazawa, Y., Ndongala, G. M., Pukuta, E., Karhemere, S., Lushima, R. S., Ilunga, B. K., Kabamba, J., Wilkins, K., Gao, J., Li, Y., Emerson, G., Damon, I. K., Carroll, D. S., Reynolds, M. G., Malekani, J., & Tamfum, J. J. M. (2015). Case report: Human monkeypox in the kivus, a conflict region of the Democratic Republic of the Congo. American Journal of Tropical Medicine and Hygiene, 93(4), 718–721. 10.4269/ajtmh.15-0095

9. Kibungu, E. M., Vakaniaki, E. H., Kinganda-Lusamaki, E., Kalonji-Mukendi, T., Pukuta, E., Hoff, N. A., Bogoch, I. I., Cevik, M., Gonsalves, G. S., Hensley, L. E., Low, N., Shaw, S. Y., Schillberg, E., Hunter, M., Lunyanga, L., Linsuke, S., Madinga, J., Peeters, M., Cigolo, C. M., . . .Research Consortium, I. M. (2024). Clade I–Associated Mpox Cases Associated with Sexual Contact, the Democratic Republic of the Congo. Emerging Infectious Diseases, 30(1), 172–176. 10.3201/eid3001.231164

10. Schwartz, D. A., & Pittman, P. R. (2023). Mpox (Monkeypox) in Pregnancy: Viral Clade Differences and Their Associations with Varying Obstetrical and Fetal Outcomes. Viruses, 15(8). 10.3390/v15081649

11. World Health Organization (WHO). Mpox (monkeypox) - Democratic Republic of the Congo. Geneva : WHO; 23 Nov 2023. Availablefrom:https://www.who.int/emergencies/disease-outbreak-news/item/2023-DON493

12. Cuérel, A., Favre, G., Vouga, M., & Pomar, L. (2022). Monkeypox and Pregnancy: Latest Updates. In Viruses (Vol. 14, Issue 11). NLM (Medline). 10.3390/v14112520

13. Schwartz, D. A., Mbala-Kingebeni, P., Patterson, K., Huggins, J. W., & Pittman, P. R. (2023). Congenital Mpox Syndrome (Clade I) in Stillborn Fetus after Placental Infection and Intrauterine Transmission, Democratic Republic of the Congo, 2008. Emerging Infectious Diseases, 29(11), 2198–2202. 10.3201/eid2911.230606

14. Low, N., Bachmann, L. H., Ogoina, D., McDonald, R., Ipekci, A. M., Quilter, L. A. S., & Cevik, M. (2023). Mpox virus and transmission through sexual contact: Defining the research agenda. PLoS Medicine, 20(1). 10.1371/journal.pmed.1004163

15. Wick, J. M., Pelliccione, A., Tran, N., & Skarbinski, J. (2024). Concurrent Sexually Transmitted Infections with Mpox Infections : A Brief Review. 10.3201/eid2704

16. Kaler, J., Hussain, A., Flores, G., Kheiri, S., & Desrosiers, D. (2022). Monkeypox: A Comprehensive Review of Transmission, Pathogenesis, and Manifestation. Cureus. 10.7759<otherinfo>/cureus.26531<lt;/otherinfo>

17. Mills, M. G., Juergens, K. B., Gov, J. P., McCormick, C. J., Sampoleo, R., Kachikis, A., Amory, J. K., Fang, F. C., Pérez-Osorio, A. C., Lieberman, N. A. P., & Greninger, A. L. (2023a). Evaluation and clinical validation of monkeypox (mpox) virus real-time PCR assays. Journal of Clinical Virology, 159. 10.1016/j.jcv.2022.10537

18. Huo, S., Chen, Y., Lu, R., Zhang, Z., Zhang, G., Zhao, L., Deng, Y., Wu, C., & Tan, W. (2022). Development of two multiplex real-time PCR assays for simultaneous detection and differentiation of monkeypox virus IIa, IIb, and I clades and the B.1 lineage. *Biosafety and Health*. 10.1016/j.bsheal.2022.10.005

19. Li, Y., Olson, V. A., Laue, T., Laker, M. T., & Damon, I. K. (2006). Detection of monkeypox virus with real-time PCR assays. Journal of Clinical Virology, 36(3), 194–203. 10.1016/j.jcv.2006.03.012

20. Kulesh, D. A., Loveless, B. M., Norwood, D., Garrison, J., Whitehouse, C. A., Hartmann, C., Mucker, E., Miller, D., Wasieloski, L. P., Huggins, J., Huhn, G., Miser, L. L., Imig, C., Martinez, M., Larsen, T., Rossi, C. A., & Ludwig, G. V. (2004). Monkeypox virus detection in rodents using real-time 3′-minor groove binder TaqMan® assays on the Roche LightCycler. Laboratory Investigation, 84(9), 1200–1208. 10.1038/labinvest.3700143

21. Chastel, C. (2009). Le monkeypox humain. In Pathologie Biologie (Vol. 57, Issue 2, pp. 175– 183). 10.1016/j.patbio.2008.02.006

22. Huang, Y., Mu, L., & Wang, W. (2022). Monkeypox: epidemiology, pathogenesis, treatment and prevention. In Signal Transduction and Targeted Therapy (Vol. 7, Issue 1). Springer Nature. 10.1038/s41392-022-01215-4

23. Damaso, C. R. (2022). The 2022 monkeypox outbreak alert: Who is carrying the burden of emerging infectious disease outbreaks? In The Lancet Regional Health - Americas (Vol. 13). Elsevier Ltd. 10.1016/j.lana.2022.100315

24. Liang, C., Qian, J., & Liu, L. (2022). Biological characteristics, biosafety prevention and control strategies for the 2022 multi-country outbreak of monkeypox. Biosafety and Health. 10.1016/j.bsheal.2022.11.001

25. Vakaniaki, E. H., Kaciat, C., Kinganda-Lusamaki, Eddy, Áine O’toole, ;, Wawina-Bokalanga, T.,Mukadi-Bamuleka, D., Aziza, A. A., Mujula, Y., Parker, ; Edyth Muswamba-Kayembe, P.-C., Nundu, S. S., Lushima, R. S., Claude, J., Cigolo, M., Mulopo-Mukanya, N., Elisabeth, ;, Simbu, P., Prince Akil-Bandali, ;, Kavunga, ; Hugo, … Placide Mbala-Kingebeni, ; (n.d.). Sustained Human Outbreak of a New MPXV Clade I Lineage in Eastern Democratic Republic of the Congo. 10.1101/2024.04.12.24305195

26. Wickham H (2016). ggplot2: Elegant Graphics for Data Analysis. Springer-Verlag New York. ISBN 978-3-319-24277-4

27. Pebesma E (2018). “Simple Features for R: Standardized Support for Spatial Vector Data.” The R Journal, 10(1), 439–446. doi:10.32614/RJ-2018-009, 10.32614/RJ-2018-009.

28. Kahle D, Wickham H. ggmap: Spatial Visualization with ggplot2 [Internet]. Vol. 5, The R Journal. 2013. p. 144–61. Available from: https://journal.r-project.org/archive/2013-1/kahle-wickham.pdf

